# Evaluation of the Socio-Medical Assessment of Work Capacity in Patients with Colorectal Cancer in German Rehabilitation Clinics: Its Diagnostic Accuracy for Actual Return to Work and the Physician’s Views on Potential Changes in Current Practice

**DOI:** 10.1101/2025.09.24.25336529

**Authors:** Tomislav Vlaski, Reiner Caspari, Harald Fischer, Tanja Trarbach, Marija Slavic, Hermann Brenner, Ben Schöttker

**Author notes:** **Corresponding author** Correspondence to Ben Schöttker. Correspondence.

## Abstract

**Simple Summary:** In Germany, physicians assess the work ability of colorectal cancer patients at the end of a usually 3-week stay in a rehabilitation clinic. This study evaluated the diagnostic accuracy of this assessment for return to work and the open-mindedness of physicians towards changing their current assessment practice. We conducted an online survey with physicians and followed up colorectal cancer patients for 9 months after rehabilitation. Although 97% of the patients were judged to be able to work by their physicians, only 70% actually did 9 months after rehabilitation. The majority of physicians at least partly agreed that a standardized checklist is needed to help them with the work ability assessment (73%), and almost all (95%) said that they would use it. The diagnostic accuracy of the current work ability assessment for colorectal cancer patients in Germany is limited, and a checklist could help physicians in their judgments.

**Background/Objectives:** Return to work (RTW) is a goal of many patients with colorectal cancer (PwCRC) attending inpatient rehabilitation. In German rehabilitation clinics, physicians conduct the socio-medical assessment of work capacity (SMWC) with the aim of assessing the current ability to work. We tested how well it also predicts the actual RTW of PwCRC.

**Methods:** This study combined a nationwide physician survey (N = 38) with longitudinal data from a cohort study of PwCRC (N = 172) aged 65 or younger who were employed prior to CRC diagnosis. Physicians were asked about their use of validated tools for the SMWC and their attitudes towards a standardized assessment checklist. PwCRC completed baseline and 9-month follow-up questionnaires about their employment status. SMWC results of the cohort study’s participants were extracted from rehabilitation discharge reports.

**Results:** While 97% of PwCRC were predicted to be capable of working **≥**6 hours/day, only 70% actually returned to work 9 months after rehabilitation. The SMWC showed high sensitivity (98%) but low specificity (6%) for predicting RTW, with a positive predictive value (PPV) of about 70%. Most physicians (73%) at least partly saw the need for an evidence-based structured checklist for an improved SMWC, and almost all (95%) would use it if it did not take more than 10 minutes to apply it.

**Conclusions:** The SMWC for PwCRC in German rehabilitation clinics is not standardized and overestimates the return-to-work rate. There is a need for a standardized checklist, and most physicians would be willing to use it.

## 1. Introduction

Colorectal cancer (CRC) is a significant global health concern, ranking as the third most frequently diagnosed malignancy and the second leading cause of cancer-related mortality internationally [1]. Survival outcomes vary widely depending on stage at diagnosis and access to healthcare, with five-year survival dropping from around 90% for localized CRC to less than 20% for metastatic cases [2, 3]. As treatment improves, CRC survivorship has become increasingly important. Many patients with CRC face challenges returning to work due to physical, psychological, and social factors, underscoring the need for supportive interventions and policies [4, 5]. Only around two thirds (58-73%) of patients with CRC return to work in the first year after rehabilitation [6-9]. For example, representative routine data from the German Pension Insurance (DRV Bund) demonstrated that 58% of patients with CRC were employed one year after rehabilitation and this proportion even slightly decreased to 53% after two years [6].

An age closely to the retirement age was the strongest predictor of unsuccessful return-to-work in the DRV Bund data analysis. Other studies showed that the most commonly reported challenges faced by patients with CRC in returning to work include physical limitations such as persistent fatigue, a lack of energy, and treatment-related complications [10, 11]. A systematic review identified that age and comorbidity, along with the nature of treatment, like the extent of surgical resection and subsequent therapies, were negatively associated with the odds of returning to work [12, 13].

Rehabilitation of cancer patients in Germany is part of a comprehensive healthcare system designed to support recovery and facilitate return to work after treatment. Hospital physicians or social workers typically initiate post-acute inpatient rehabilitation, whereas rehabilitation not following acute treatments can be requested directly by patients with supporting statements of their physicians. Inpatient rehabilitation in Germany typically lasts three weeks and is initiated by hospital physicians or social workers. The programs in oncological rehabilitation adopt a holistic, interdisciplinary approach— combining physical therapy, occupational counseling, patient education, and psychosocial support—to help patients adapt to life post-cancer and improve coping mechanisms [14]. Return to work (RTW) is a central goal, given its positive impact on quality of life [15]. Although participation in rehabilitation is a statutory right, studies show that uptake remains suboptimal, with only 60–70% of eligible patients participating [16, 17]. Barriers include logistical issues, limited awareness, and socio-economic disparities.

Multidisciplinary care, especially when combining psychoeducational, vocational, and physical interventions, has been shown to improve return-to-work outcomes [18-20].

In Germany, a key component of the rehabilitation process is the “*Sozialmedizinische Leistungsbeurteilung”*, which can be translated as “socio-medical assessment of work capacity” (SMWC) [21]. Conducted by a physician at the end of a rehabilitation program, the SMWC evaluates a patient’s functional abilities and capacity to participate in work and daily activities. Its primary purpose is to assess immediate work ability at the time of discharge, not to predict long-term return-to-work outcomes.

Nevertheless, the SMWC is crucial for determining eligibility for social benefits and a disability pension by the German Pension Insurance. While the SMWC is a mandatory component of the rehabilitation process in Germany and serves as a basis for determining work capacity and entitlement to social benefits, there is a lack of data on its predictive validity for actual return to work at later time points and on how it is being conducted in practice. As no standardized instrument currently guides the SMWC, a large variability in its outcomes can be expected, and it is of interest whether physicians in rehabilitation clinics would be open-minded towards using a standardized checklist to help them with the SMWC

In a CRC patient cohort study, we aimed to evaluate the diagnostic accuracy of the SMWC conducted during inpatient rehabilitation in predicting return to work nine months after rehabilitation. In addition, we conducted a physician survey on the current practice of SMWC in Germany and on their perspectives on potential changes, like the potential implementation of a standardized checklist.

## 2. Materials and Methods

Two distinct studies were conducted to address the aims of this project: A cross-sectional physician survey and a cohort study with patients with CRC.

### 2.1 Physician Survey

A cross-sectional survey was conducted among physicians involved in the rehabilitation of patients with CRC across Germany. The head physicians of the oncological wards of all 62 rehabilitation clinics offering oncologic rehabilitation for patients with CRC in Germany were sent the invitation to participate in an online survey via regular mail, and the freely available mail addresses on the internet homepages of the clinics were used. The invitation letter asked the head physicians to distribute the invitations to the online survey to the whole team of physicians conducting SMWCs for patients with CRC to ensure participation of physicians from all career stages. The first mail was sent on December 18, 2024, and a reminder was sent on January 12, 2025.

The online survey contained 15 questions, divided into three segments:

1. Professional experience: Questions about years of experience, specialization, frequency of performing SMWCs, and time spent on each assessment.
2. SMWC in practice: Questions regarding the physicians’ perception of their own performance in predicting the return to work of patients with CRC, the criteria used for assessment, and whether validated tests were employed.
3. Acceptance of the implementation of a novel checklist for SMWCs: Questions about the physicians’ views on the need for a checklist for SMWCs, their willingness to use it, potential barriers, and required support measures.

### 2.2 Cohort of patients with CRC

The participants for this analysis were selected from the ongoing MIRANDA study, a multicenter, prospective cohort project focusing on fatigue, quality of life, return to work, and other health-related outcomes in patients with CRC undergoing in-patient rehabilitation (registration number:

DRKS00020822) [22]. Participants in this study were recruited in 6 centers located in all parts of Germany. Inclusion criteria were: an age of 18 years and older, a CRC diagnosis, relevant cancer treatment (surgery, chemotherapy, and/or radiotherapy) in the past 12 months, and a planned in-patient rehabilitation for at least 3 weeks. For this specific project, we only included patients with CRC, who should be available on the labor market 9 months after oncological rehabilitation. Thus, we excluded MIRANDA study participants who were not employed before CRC diagnosis, were older than 65 years during rehabilitation. Furthermore, we excluded participants who had a CRC diagnosis more than 12 months before the baseline assessment in the rehabilitation clinics.

In total, 616 patients with CRC got enrolled between Sept 2020 and April 2024 in the MIRANDA study and had completed the 9-month follow-up at the time of this analysis (Spring 2025). Of these, 384 did not meet the eligibility criteria outlined above, leaving 232 participants eligible for this project. Overall, 189 of these 232 participants (81.5%) participated in the 9-month follow-up. Further 17 study participants needed to be excluded due to missing data, leaving 172 patients with CRC for analysis (**Figure 1**).

**Figure 1.**
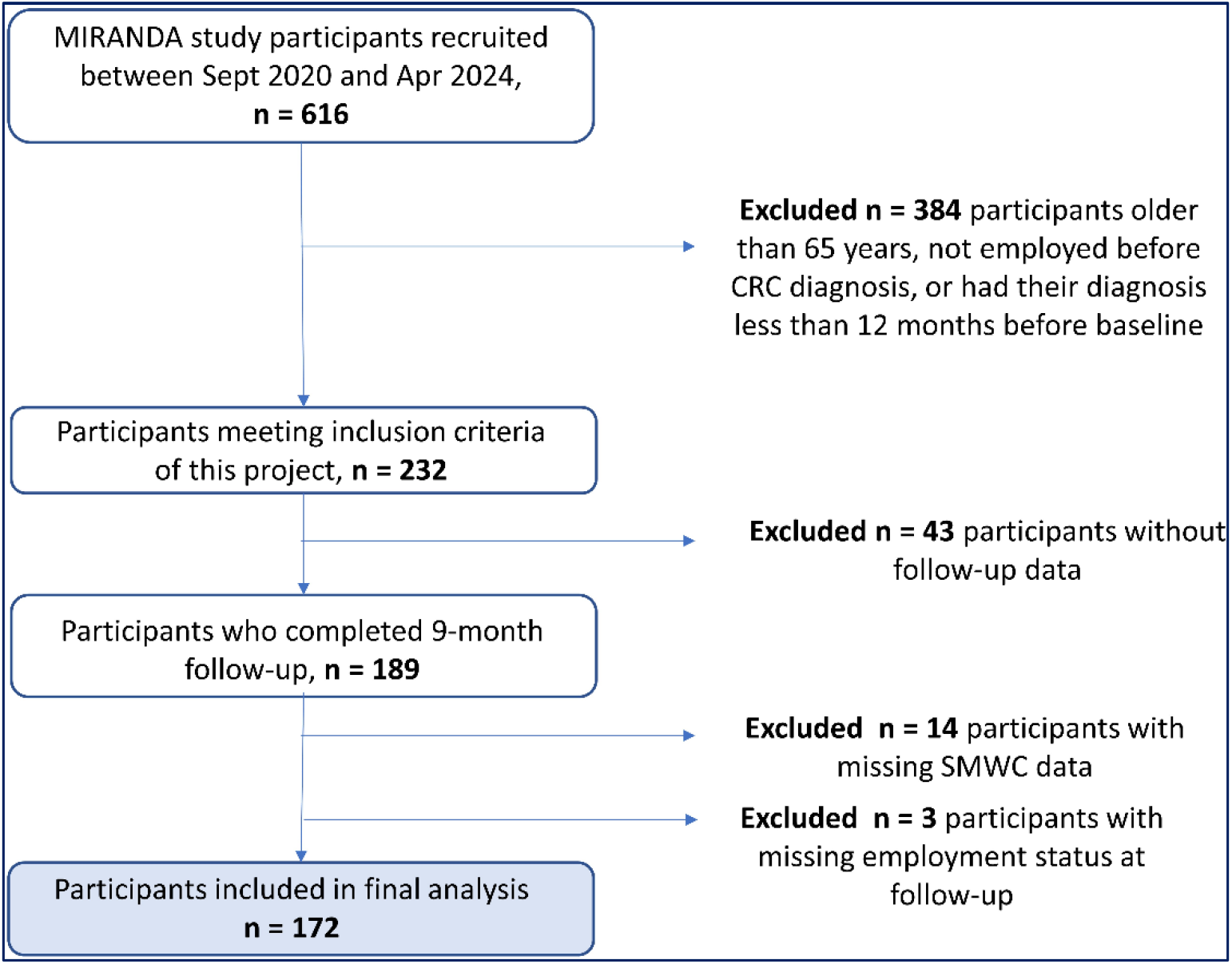
Flow chart of MIRANDA study participants who should be available on the labor market 9 months after oncological rehabilitation and could be included in the final analysis of this project.

Data on sociodemographic information (e.g., age, sex), lifestyle factors (e.g., physical activity levels, smoking status, BMI), clinical data (e.g., cancer treatment, comorbidities), and the employment status prior CRC diagnosis (incl. total number of paid working hours per week) were collected from patients with CRC through baseline questionnaires administered during rehabilitation. Weight and height were measured, and cancer stage was physician-reported.

The SMWC was performed at the end of the rehabilitation stay by the attending physicians and documented in the medical discharge report, from which it was extracted at a later stage of the project.

The assessment is based on a structured evaluation of each patient’s qualitative functional capacities, including both positive resources (e.g., physical endurance, lifting/carrying ability, mobility, concentration, and psychological resilience) and negative limitations (e.g., fatigue, pain, cognitive impairments, and stoma-related restrictions). These qualitative judgements are then summarized into a quantitative performance profile, which categorizes the expected number of working hours a patient can perform per day. In line with German Pension Insurance guidelines, physicians assign patients to one of three categories:

- 6 hours or more
- Between 3 and 6 hours
- 3 hours or less

This categorization is documented separately for the general job market and for the previous occupation, which may involve specific physical or organizational requirements.

At the 9-month follow-up, participants completed standardized questionnaires that included two items on their current employment status. The first item asked participants to indicate their current work status, with five response options:

a. Not working due to CRC
b. Not working for other reasons
c. Working fewer hours than before the CRC diagnosis due to the disease
d. Working fewer hours than before the CRC diagnosis for other reasons
e. Working the same or more hours than before the CRC diagnosis

The second item, applicable to those currently employed, asked for the total number of paid working hours per week. For analysis, responses (a) and (b) were categorized as “Unsuccessful return to work,” while responses (d) and (e) were categorized as “Successful return to work”. Participants who selected option (c)—working fewer hours because of CRC—were further classified based on their reported working hours per day: those working at least three hours per day were assigned to the “successful” group, and those working fewer than three hours per day were assigned to the “unsuccessful” group.

Moreover, the work ability index (WAI) was administered at the 9-month follow-up to quantitatively assess the work performance capacity of those CRC survivors who returned to work [23]. The WAI is a validated self-report instrument that evaluates an individual’s ability to work in relation to their health status and the specific demands of their job. It has several dimensions, including current work ability compared to one’s lifetime best, the impact of health conditions on work performance, the number of diagnosed diseases, recent sick leave, and personal expectations regarding future work ability. Each component is scored, and the overall score is calculated by summing these individual scores to yield a total score ranging from 7 to 49. Based on the total score, work ability is categorized into four levels: poor (7–27), moderate (28–36), good (37–43), and excellent (44–49).

### 2.3 Statistical analysis

All analyses were done using SAS software version 9.4. Descriptive statistical measures were used to characterize the results of the physicians’ survey and the baseline characteristics of the CRC patient cohort.

For the SMWC, the three original categories (“6 hours or more”, “from 3 to 6 hours”, and “under 3 hours”) were dichotomized into “Sufficient work ability” (≥3 hours) versus “Insufficient work ability” (<3 hours). The return-to-work outcome, measured at the 9-month follow-up, was also dichotomized as “successful” or “unsuccessful.” Two contingency tables of return to work status and SMWC were created, one for work ability prediction for the general job market and the other for work ability prediction for the previous job. Accuracy, sensitivity, specificity, positive predictive value (PPV), and negative predictive value (NPV) with 95% confidence intervals were calculated. All analyses were conducted as a complete-case analysis.

## 3. Results

### 3.1. Physician Survey

A total of 38 physicians participated in the survey, and their responses about their professional experience and training are shown in **Table 1**. A fifth of the survey participants had less than 5 years and another fifth 5–15 years of experience with SMWCs. Most physicians had between 16 and 25 years of professional experience (40%), while 8% reported a longer time. The most commonly reported specialties were internal medicine (47%) and oncology (29%), followed by general medicine (24 %), gastroenterology (1 %), physical and rehabilitative medicine (13 %), and psychosomatic medicine and psychotherapy ( 7%). These numbers do not add up to 100 % because several physicians had two or more specialties. Only n=7 participants held no medical specialty (15%). Approximately one-third of respondents (34%) indicated they had participated in advanced training in social medicine. Regarding the frequency of SMWCs, 90% conducted them almost every week, while the remaining 10% did so at least once a month. Around half of the physicians (51%) reported spending between 15 and 30 minutes per assessment, while 41% completed them in under 15 minutes and 8% required more than 30 minutes. The median time per assessment was 15 minutes (interquartile range: 10–30 minutes).

**Table 1.** Professional experience and education level of the participants of the physician survey.

| Physician data | N<sub>total</sub> | Proportion (%) | Mean (95%CI) |
| --- | --- | --- | --- |
| Years of professional experience in conducting socio-medical assessments of work capacity (SMWCs) | 38 |  | 15 (12 – 18) |
| <5 |  | 10 (26) |  |
| 5 - 15 |  | 10 (26) |  |
| 16 - 25 |  | 15 (40) |  |
| >25 |  | 3 (8) |  |
| Specialties held | 38 |  |  |
| Internal medicine |  | 18 (47) |  |
| Oncology |  | 11 (29) |  |
| General medicine |  | 9 (24) |  |
| Gastroenterology |  | 6 (16) |  |
| Physical & rehabilitative medicine |  | 5 (13) |  |
| Psychosomatic medicine & psychotherapy |  | 1 (7) |  |
| Advanced training in social medicine | 38 |  |  |
| Yes |  | 13 (34) |  |
| No |  | 25 (66) |  |
| Frequency of conducting SMWCs | 38 |  |  |
| Almost every week |  | 34 (90) |  |
| At least once a month |  | 4 (10) |  |
| Minutes per SMWC | 37 |  | 21 (16 – 25) |
| <15 |  | 15 (41) |  |
| 15 - 30 |  | 19 (51) |  |
| 30> |  | 3 (8) |  |

**Table 2** shows the responses of the physicians about the SMWCs in practice. When asked about the criteria used for the SMWC, most physicians reported considering both the physical and mental demands of the patient’s current job (n = 35 (92%) each). Other common factors considered included the general physical fitness (n = 33, 87%), cancer stage (n = 32, 84%), comorbidities (n = 29, 76%), and the presence of a stoma (n = 29, 76%). Less frequently mentioned factors involved chronic pain (n = 24, 63%), age (n = 17, 45%), and sleep disturbances (n = 16, 42%). Only half of the respondents (n = 19, 50%) reported using validated instruments for the SMWC. Among these, 16 physicians provided further information on the domains of the tests applied. The most frequently mentioned domain was physical performance (n = 9; e.g., 5- or 6-minute walk test, ergometer, hand grip strength), followed by pain and symptoms (n = 6; e.g., Visual Analog Scale, neuropathy or pain questionnaires), mental health (n = 6; e.g., Hospital Anxiety and Depression Scale, PHQ-4, psycho-oncological testing), and fatigue or distress (n = 5; e.g., Brief Fatigue Inventory, Distress Thermometer). In addition, some physicians mentioned tools from other areas, such as occupational therapy workplace assessments, body composition diagnostics, or organ-specific tests (e.g., spirometry, ENT diagnostics).

**Table 2.** Questions about how socio-medical assessments of work capacity (SMWC) are being performed by physicians in practice in oncologic rehabilitation clinics in Germany for colorectal cancer patients and on the acceptance of a validated checklist to support physicians in the SMWC.

| Question | N<sub>total</sub> | Proportion (%) | Mean (95%CI) |
| --- | --- | --- | --- |
| Criteria used for the SMWC | 37 |  |  |
| Physical demands of the current job |  | 35 (95) |  |
| Mental demands of the current job |  | 35 (94) |  |
| General physical fitness |  | 33 (89) |  |
| Cancer stage |  | 32 (87) |  |
| Chemo- and/or radiotherapy |  | 32 (87) |  |
| Nutritional status |  | 31 (84) |  |
| Comorbidities |  | 29 (78) |  |
| Stoma present |  | 29 (78) |  |
| Chronic pain |  | 24 (64) |  |
| Age |  | 17 (46) |  |
| Sleep disturbances |  | 16 (43) |  |
| Weight |  | 9 (24) |  |
| Education level |  | 6 (16) |  |
| Gender |  | 2 (5) |  |
| Use of validated tests | 38 |  |  |
| Yes |  | 19 (50) |  |
| No |  | 19 (50) |  |
| Domains of the validated test used: | 16 |  |  |
| Physical performance |  | 9 (24) |  |
| Functional/occupational |  | 1 (3) |  |
| Mental health |  | 6 (16) |  |
| Fatigue or distress |  | 5 (13) |  |
| Pain and other subjective symptoms |  | 6 (16) |  |
| Body composition diagnostics |  | 2 (5) |  |
| Other |  | 2 (5) |  |
| Estimation of validity of SMWCs in practice |  |  |  |
| Percentage correctly assessed as able to work | 37 |  | 68 (62– 74) |
| Percentage correctly assessed as unable to work | 37 |  | 79 (70 – 88) |
| Support measures needed to improve SMWC quality | 37 |  |  |
| More time resources |  | 5 (14) |  |
| Training |  | 7 (19) |  |
| Access to standardized assessment instruments |  | 24 (65) |  |
| More interdisciplinary collaboration |  | 11 (30) |  |
| No support measures needed |  | 9 (24) |  |
| Agreement with the need of a validated checklist | 37 |  |  |
| Somewhat/strongly agree |  | 15 (40.5) |  |
| Partly agree/partly disagree |  | 12 (32.4) |  |
| Somewhat/strongly disagree |  | 10 (27.0) |  |
| Willingness to use a checklist if it takes 10 min. to apply it | 37 |  |  |
| Yes |  | 34 (91.9) |  |
| No |  | 3 (8.1) |  |

| Question | N<sub>total</sub> | Proportion<br>(%) | Mean<br>(95%CI) |
| --- | --- | --- | --- |
| Agreement with statements about use of a validated checklist | 37 |  |  |
| A standardized checklist would lead to more accurate assessments |  | 17 (45.9) |  |
| A standardized checklist would increase time efficiency |  | 11 (29.7) |  |
| A standardized checklist would lead to better documentation |  | 22 (59.5) |  |
| A standardized checklist would lead to more comparable assessments among different physicians |  | 30 (81.1) |  |
| Practical barriers for implementation of a checklist | 37 |  |  |
| Yes |  | 13 (35.1) |  |
| No |  | 24 (64.9) |  |
| If yes, which barriers? | 13 |  |  |
| Not enough time to use it |  | 2 (5) |  |
| Lack of acceptance among colleagues |  | 4 (11) |  |
| Would be perceived as additional bureaucracy |  | 9 (24) |  |
| A training would be required |  | 5 (14) |  |

The physicians were further asked to estimate, in percentage, how many of patients with CRC they judged to be ‘able to work’ will actually return to work. The mean estimate was 68% (95% confidence interval (CI): 62 - 74). Additionally, they were asked to estimate how many of the patients they labeled as ‘unable to work’ would actually not return to work. The mean (95% CI) estimate for this group was 90% (80-95). When asked about support measures they would need to improve the quality of the SMWCs, the access to standardized assessment instruments (64.9%) was the most commonly named measure. Other support measures were only needed by a minority of the physicians, and every 4^th^ physician (24%) even replied that they don’t need support at all.

**Table 2** also shows the responses from 37 physicians about their perception of a validated checklist to support physicians in conducting SMWCs for patients with CRC. In terms of agreement with the need for such a checklist, the majority agreed (15, 41%) but there were also many physicians who partly agreed/partly disagreed (12, 32%) or disagreed (10, 27%). However, on the condition that the use of such a checklist would not require more than 10 minutes of their time, almost all respondents would use it in practice (34, 92%). When considering statements about the potential benefits of a standardized checklist, 45.9% believed it would lead to more accurate assessments, 29.7% expected increased time efficiency, 59.5% anticipated improved documentation, and 81.1% said it would result in more comparable assessments among different physicians. When asked about barriers for the implementation of a checklist in practice, 35% of respondents acknowledged issues, with the most frequently stated barrier being that the checklist would be perceived as additional bureaucracy (9, 24%). Other barriers were rarely stated. Most importantly, only two respondents (5%) said that there would not be enough time in practice to use it.

### 3.2. Colorectal Cancer Patient Cohort

#### 3.2.1. Baseline description

**Table 3** shows the baseline characteristics of the 172 patients included in the CRC patient cohort. The median age was 56 years (IQR, 52–60), with nearly two-thirds (60.5%) being male and approximately 39.5% female. The majority (58.1%) had a tumor in the colon, fewer participants in the rectum (36.1%), and a minority (5.8%) in both colon and rectum. While CRC stages I–III were approximately equally distributed at 32.6%, 31.4%, and 30.2%, respectively, only 4.6% of patients had stage IV disease. Nearly all patients underwent surgery (98.8%), just over half received chemotherapy (52.4%), and approximately one in five received radiotherapy (18.3%). The median BMI was 26.9 kg/m^2^ (IQR, 23.5–30.5), and almost two-thirds of the patients (66.3%) were overweight or obese. Regarding smoking status, 44.7% had never smoked, 40.6% were former smokers, and 14.7% were current smokers. Less than half of the patients (48.5%) reported engaging in a healthy level of physical activity. Approximately every 4^th^ study participant was free of comorbidity (27.2%). Most participants had one comorbidity (32.5%), followed by no comorbidities (27.2%), and two comorbidities (23.1%). The most common comorbidities were hypertension (41.4%) and diabetes mellitus (11.2%). Most patients were fully employed (82.0%), while the remainder worked part-time (18.0%).

**Table 3.**
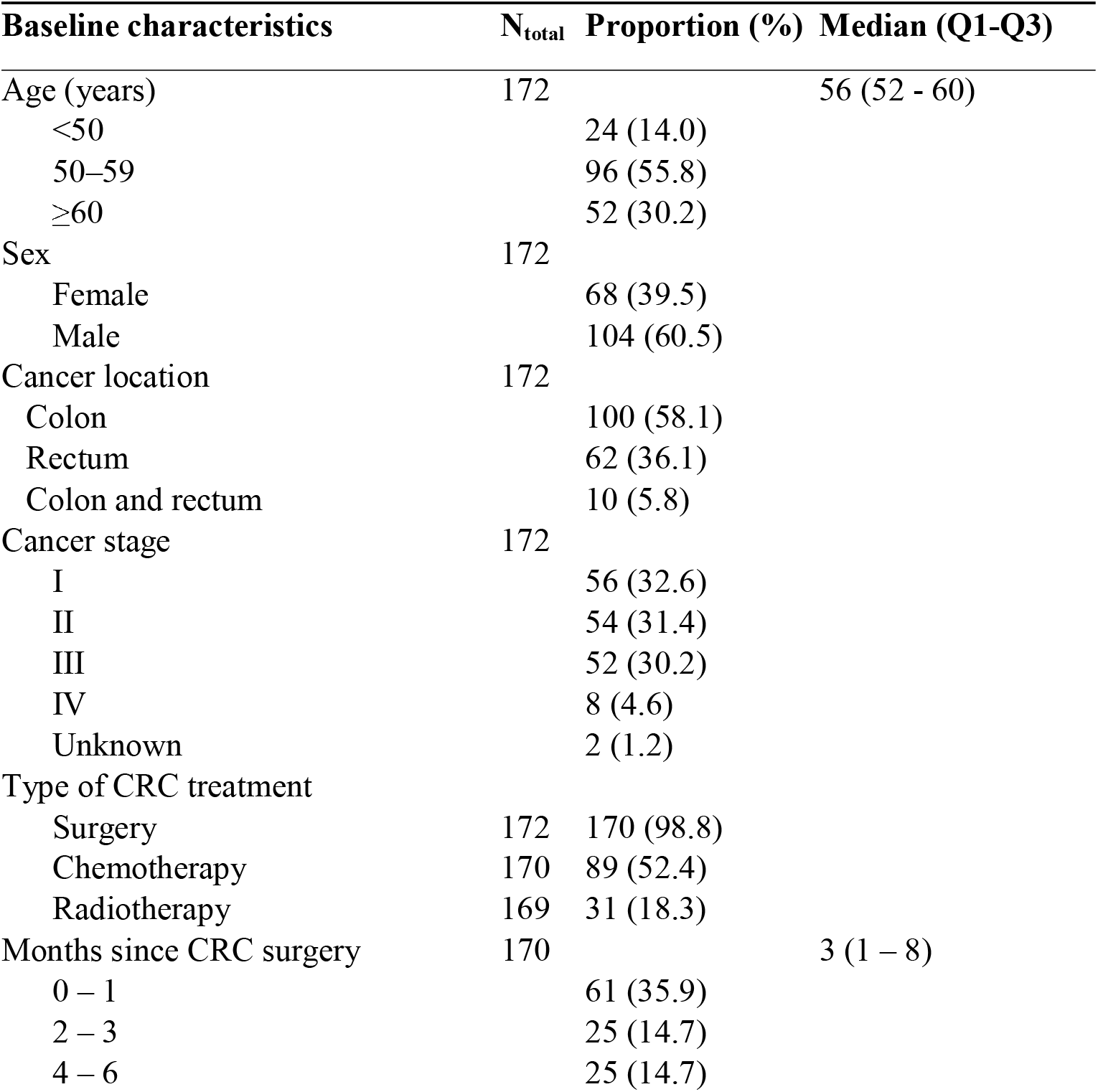
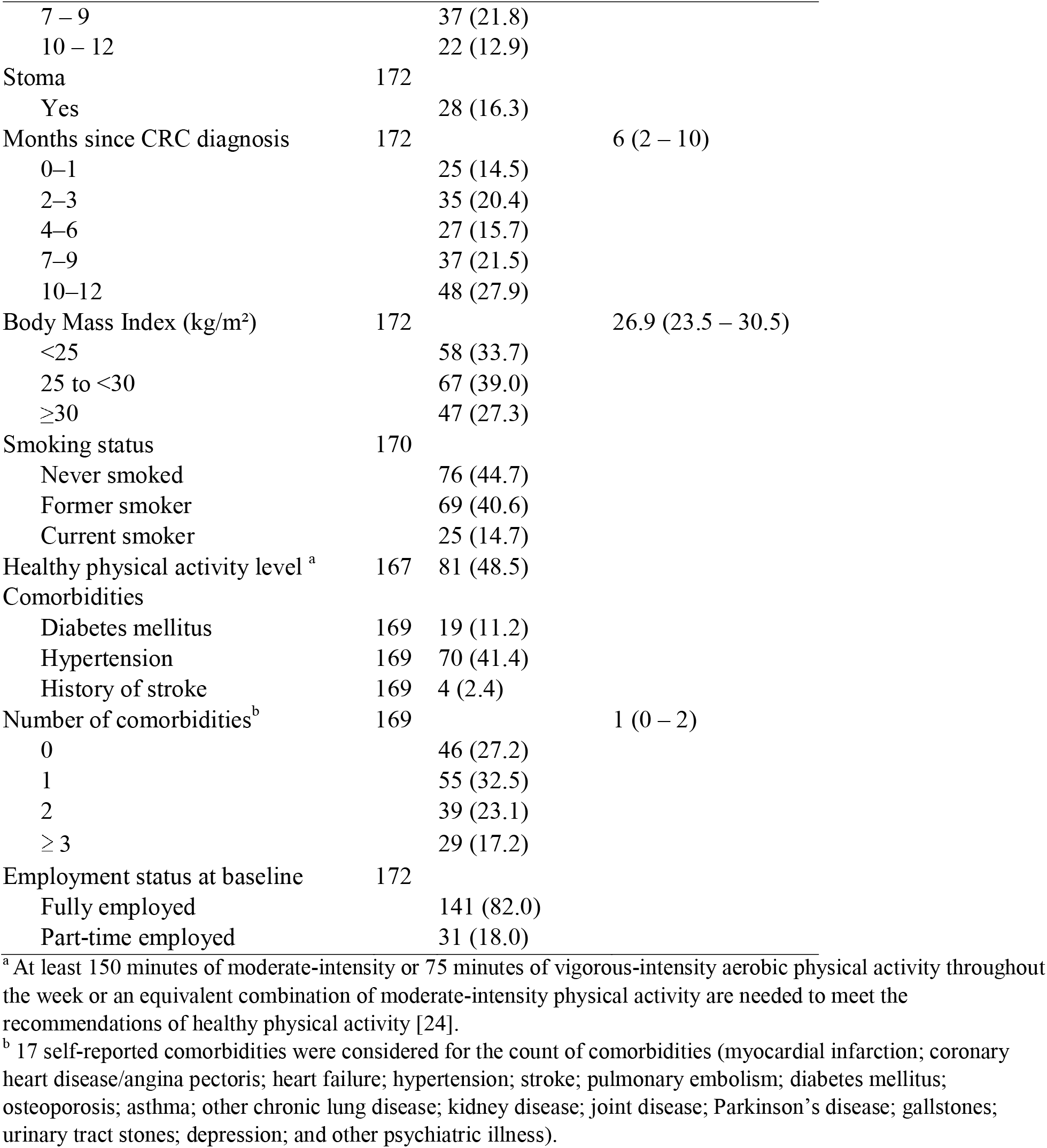
Baseline characteristics of the colorectal cancer patient cohort.

#### 3.2.2. Diagnostic Accuracy of the Socio-medical assessment of work capacity for Actual Return to Work

Among the 172 patients with available data, 121 (70.3%) returned to work within 9 months (**Table 4**). About 7 out of 10 of those patients with CRC classified as having sufficient work capacity for the general job market (71%) or the previous job (73%) by the SMWC were in fact working 9 months after oncologic rehabilitation. In contrast, only about 6 out of 10 of those judged to have insufficient capacity for the general job market (60%) or the previous job (62%) did not return to work.

**Table 4.** Contingency table of work ability prediction by the socio-medical assessment of work capacity (SMWC) for the general job market and the previous job in relation to actual return to work 9 months after rehabilitation.

| Work ability assessment for the general job market | Status of actual return to work |  |  | Work ability assessment for the previous job | Status of actual return to work |  |  |
| --- | --- | --- | --- | --- | --- | --- | --- |
|  | Successful return to work | Unsuccessful return to work | Total |  | Successful return to work | Unsuccessful return to work | Total |
| Sufficient work ability | 119 | 48 | 167 | Sufficient work ability | 116 | 43 | 159 |
| Insufficient work ability | 2 | 3 | 5 | Insufficient work ability | 5 | 8 | 13 |
|  | 121 | 51 | 172 |  | 121 | 51 | 172 |

**Table 5** summarizes the diagnostic accuracy of the work ability assessment by the SMWC for return to work prediction. The SMWC demonstrated high sensitivities (>95%) and low specificities (<16%) for the prediction of the actual return to work in the previous job or to get employed in the general work market. The positive predictive values (PPV) were around 70%, and the negative predictive values (NPV) were close to 60%. In summary, the accuracy of the SMWC for prediction of employment in any job and return to work to the previous job were 71% and 72%, respectively.

**Table 5.** Diagnostic accuracy of the socio-medical assessment of work capacity (SMWC) at the end of rehabilitation for the actual return to work.

| Dimension of the SMWC | Sensitivity (%)<br>(95% CI) | Specificity (%)<br>(95% CI) | Positive predictive value (%)<br>(95% CI) | Negative predictive value (%)<br>(95% CI) | Accuracy (%)<br>(95% CI) |
| --- | --- | --- | --- | --- | --- |
| Work ability prediction for the general job market | 98.3<br>(94.2–99.5) | 5.9<br>(2.0–15.9) | 71.3<br>(64.0– 77.6) | 60.0<br>(23.1–77.6) | 70.9<br>(63.7 – 77.2) |
| Work ability prediction for the previous job | 95.9<br>(90.7–98.2) | 15.7<br>(8.2–28.0) | 73.0<br>(65.6 – 79.3) | 61.5<br>(35.5–82.3) | 72.1<br>(64.0 – 77.7) |

#### 3.2.3. Work ability of colorectal cancer patients who returned to work

Of the 172 study participants employed before CRC diagnosis, 121 (70%) returned to work 9 months after rehabilitation. Among these, 111 provided complete Work Ability Index (WAI) data. The majority rated their work ability as moderate or poor (67%). The mean (95% CI) total WAI score was 32 points (30–34), which falls into the “moderate work ability” category (28–36 points).

A comparison of the employment status before CRC diagnosis with the employment status 9 months after rehabilitation showed that of those 121 participants who returned to work, 102 had been employed full-time before diagnosis. Of these, 75 (74%) continued in full-time employment, while 27 (26%) shifted to part-time positions. All 19 participants who had been employed part-time before diagnosis remained in part-time employment contracts.

## 4. Discussion

### 4.1 Summary of Main Findings

The survey of 39 physicians revealed that the SMWC is typically done in 15 minutes. Physicians considered physical and mental job requirements, comorbidities, cancer stage, and overall physical condition in their assessments, but only about half used validated instruments, and the instruments used varied strongly. The majority of the physicians (73%) at least partly agreed that there is a need for a validated, standardized checklist for the SMWC, and almost all would use it (92%) on the condition that it does not take more than 10 minutes to apply it. In the colorectal cancer patient cohort, 97% of patients with CRC were judged to be capable of working ≥6 hours/day by the SMWC, but only 70% actually returned to work 9 months after rehabilitation. Performance evaluation of the SMWC showed a high sensitivity (>95%), low specificity, and moderate overall accuracy (71%) for predicting return to work. Among those who actually returned to work, the work ability was moderate or poor for three-quarters of the study population. Nevertheless, 3 out of 4 full-time employed patients with CRC who returned to work remained full-time employed, while only 1 out of 4 continued to work part-time.

### 4.2 Return-to-Work Rates and Work Ability Index Scores in other studies

A 70% return-to-work rate in our study at nine months after rehabilitation aligns with previous research. For example, a study from the Netherlands reported that 67.5% of patients with CRC resumed employment within two years, and an Australian study found that 73% of middle-aged survivors had returned to work by 12 months [7, 13]. Additionally, a study from the United Kingdom noted that 61% of patients with CRC resumed work one year after surgery, and a Danish study documented employment in 62% of patients one year after diagnosis [8, 9]. Routine data from the German Pension Insurance showed that one year after oncological rehabilitation, 58% of patients with CRC were employed [6]. Although these studies differ in follow-up periods between 1 and 2 years and patient characteristics, the 70% return-to-work rate at nine months after CRC surgery observed in this study fits within the reported range of 58%-73% by these other studies, suggesting that approximately two-thirds of CRC survivors successfully reintegrate into the workforce within the first two years after treatment.

Despite the relatively high return-to-work rate at the 9-month follow-up, our CRC cohort demonstrated a mean Work Ability Index (WAI) of 33 points, which is in the range of the “moderate work ability” category of the WAI (28–36 points) and notably lower than the mean score of 40 reported for the general population and slightly below the mean score of 36 observed among young cancer survivors [25, 26]. This low WAI suggests that, although a substantial proportion of CRC survivors resume employment, they continue to experience reduced functional capacity compared to their work colleagues of comparable age who are not cancer survivors. Moreover, the low WAI in our study aligns with evidence indicating that cancer survivors often require workplace adaptations and ongoing rehabilitation to maintain sustainable employment [27]. This can also explain why one quarter of all previously full-time employed patients with CRC chose to return to work after rehabilitation with part-time employment only. Future research should focus on identifying the specific barriers to full recovery in work ability and developing targeted interventions to support the return to work of patients with CRC.

### 4.3 Evaluation of the SMWC for return to work prediction

A total of 97% of patients were judged able to work at least 6 hours per day in any job from the general job market, compared to 92% in their previous job, which might have had specific requirements the person can no longer handle. These specific physical and organizational demands of the previous job are considered in the SMWC for that job, but not in the one for the general job market.

It should be emphasized that the primary purpose of the SMWC is to assess a patient’s ability to work after discharge from the rehabilitation clinic, not to predict their actual return-to-work status at later time points. However, we were interested in how well this ability-to-work assessment predicts actual return to work 9 months after rehabilitation discharge. The relatively long follow-up period was chosen because we wanted to ensure that all patients with CRC completed their primary cancer treatment, which often involves several cycles of chemotherapy and sometimes radiotherapy after rehabilitation.

The diagnostic accuracy of the SMWC for actual return to work was characterized by very high sensitivities (98% for the general job market and 96% for the previous job) and very low specificities (6% for the general job market and 16% for the previous job). This demonstrates a strong ability of the SMWC to correctly identify patients likely to return to work (sensitivity), while the SMWC is weak at correctly identifying patients unlikely to return (specificity). These false-positive SMWCs have practical consequences for the patients with CRC. Patients labeled as “fit for work” may face reduced access to disability pensions or other support measures helping work reintegration, and may feel pressured to resume work despite ongoing limitations. Employers may not provide the necessary job flexibility or part-time employment, making work reintegration harder. Therefore, enhancing the specificity of the SMWC is important for a fair and patient-centered decision-making in the German rehabilitation setting.

In the physician survey, physicians correctly rated that only about 70% of patients with CRC they judged to be capable of working after rehabilitation discharge will actually return to work. However, they overestimated the agreement rate of patients judged not to be capable of working who would actually not return: instead of 90%, only 60% of patients with CRC judged unable to work truly did not return to work 9 months after rehabilitation. Since we only had 5 patients with CRC in our study whom physicians rated as unable to return to work, this estimate should be interpreted with caution. We interpret this part of our study as follows: Physicians in the German oncological rehabilitation setting are aware that the SMWC has limited diagnostic accuracy regarding actual return to work after rehabilitation.

Another important question is why substantially more patients with CRC are classified as fit for the general labor market in the SMWC than are observed to return to work in practice. This overestimation could have several reasons. In the German pension insurance guidelines for the SMWC, the assessment for the general labor market is based on a standard that requires the rehabilitant to be capable of performing at least light physical work for six hours per day or more [28]. This threshold is relatively low. Therefore, patients with low to moderate ability to work need to be classified as capable of working according to the guideline. Some patients with CRC might not resume work if they can only work for just over 15 hours per week because the financial benefit from such limited employment is low or social circumstances prevent their return to previous employment [29, 30]. Additionally, it might be difficult for them to find an employer offering a job that can be done with such few hours, especially if their previous job isn’t suitable for part-time employment with limited hours. Moreover, in Germany, patients with CRC aged 62 or older can apply for early retirement through the German Pension Insurance. Another possible reason is that the progression of chemotherapy-induced polyneuropathy (CIPN), which often impacts work ability, is hard to predict. Other factors, such as tumor recurrence, challenges in reintegrating individuals with a permanent stoma or incontinence, and the emergence of other, sometimes unpredictable, comorbidities (e.g., orthopedic, cardiovascular, or psychiatric conditions), may also reduce the predictive value of the SMWC. Finally, the absence of a standardized tool for assessing work ability could be another explanation. This situation forces physicians to rely on their individual experience and interpretation of assessment tools, which are not specifically validated for the SMWC. The physician survey revealed that only half of the physicians use standardized assessment tools for the SMWC. Examples, occasionally used in the SMWC, include the Hospital Anxiety and Depression Scale (HADS) and physical performance tests, like the 6-Minute Walk Test. These tools evaluate aspects of work ability, such as physical endurance, mobility, symptom burden (e.g., fatigue, pain), or psychological well-being.

Besides methodological differences, variations in SMWC outcomes may also depend on how physicians make their judgments. Clinical decision-making often uses heuristic approaches and can be affected by cognitive biases. For instance, optimism bias might lead physicians to believe that patients who seem clinically stable at discharge will be able to take up employment. Similarly, relying on past experience instead of structured criteria can increase subjective variability. These tendencies in subjective decision-making may contribute to systematic overestimation of work ability and underscore the importance of standardized, evidence-based tools.

Psychosocial stressors also play a crucial role in successful return to work, and their long-term courses are difficult to evaluate during rehabilitation in the SMWC. Previous studies indicated that fatigue, emotional distress, and low self-confidence can impede vocational reintegration [31, 32]. Similar psychosocial stressors likely affect patients with CRC, especially those dealing with treatment side effects such as stoma-related complications, which have been linked to a decreased likelihood of returning to work below 30% [33, 34]. In our study, even among patients who did return to work, two-thirds had moderate or poor WAI scores 9 months after rehabilitation, indicating ongoing impairments in work capacity. The reasons they continue working are probably financial pressures and social isolation, which are serving as motivating factors to re-enter the workforce because they are perceived as more distressing than the physical effects of cancer [35].

### 4.4 Need for improvement of the SMWC

Although the SMWC’s purpose is to evaluate ability to work immediately after inpatient rehabilitation discharge and not to forecast return to work months later, our findings suggest that the SMWC could benefit from a standardized, validated checklist that covers all relevant aspects for a successful return to work.

This tool could be a checklist with an easy scoring system to avoid time-consuming tests. While physicians showed a strong willingness to use such a checklist, it is evident that the roughly 15 minutes they typically spend on a SMWC should not be extended. Otherwise, the tool might be seen as an unnecessary bureaucratic burden, as survey participants expressed skepticism that a validated checklist would improve the time efficiency of conducting SMWCs. The physicians were more optimistic that such a checklist would lead to better documentation and more consistent assessments among different physicians.

To ensure that such a tool is accepted and practical in routine care, future research should involve physicians and patients in its development. A structured multi-stage Delphi process would be an appropriate method to incorporate the expertise of rehabilitation physicians, reach consensus about the content of the checklist, and balance diagnostic accuracy with practicality. The International Classification of Functioning, Disability and Health Core Set for vocational rehabilitation provides a common framework to describe functioning and work-related limitations, and has recently been validated in cancer survivors in Italy [36]. In parallel, Functional Capacity Evaluation protocols offer a structured, performance-based assessment of an individual’s ability to meet job demands and are widely used internationally as part of return-to-work evaluations [37]. Building on these existing international concepts, developing and validating a tool to improve or extend the SMWC in the German rehabilitation setting would be desirable.

### 4.5 Strengths and limitations

A key strength of this study is its multidimensional design, which combines data from the physician survey and the patient-reported data from the prospective CRC patient cohort to provide a comprehensive view of work reintegration among CRC survivors. The inclusion of 172 patients with CRC recruited by five rehabilitation clinics located in different regions of Germany enhances the representativeness of our findings. However, generalizability to all patients with CRC in Germany should still be interpreted with caution. Likewise, the high response of 38 participants in the first nationwide physician survey among physicians conducting SMWCs among patients with CRC enhances the generalizability of findings within the German rehabilitation setting. As there are only 62 rehabilitation clinics offering oncologic rehabilitation for patients with CRC in Germany, there are not many potential participants in the survey (maybe approx. n=150). The Work Ability Index (WAI) helped to standardize the assessment of work ability.

Our study also has limitations. First the patient-reported employment status was not validated through employer or administrative records. Second, the nine-month follow-up period may not fully capture long-term employment patterns or delayed work resumption. However, analyses of routine data from Germany have shown that employments rates of CRC patients after rehabilitation are stable from 6 months to 2 years after rehabilitation [6]. Third, sample size limitations precluded subgroup analyses of high interest, such as those for sex and CRC stage. Finally, patient perspectives on barriers and motivations for resuming work were not captured, which limits the contextualization of the quantitative findings and highlights the need for future studies that integrate qualitative data.

## 5. Conclusions

The ability of the SMWC to predict the return to work of patients with CRC 9 months after a stay in a German rehabilitation clinic is limited due to a low specificity. This can be primarily explained by the aim of the SMWC, which is to assess the ability to conduct at least light physical work for 6 hours or more per day right after inpatient rehabilitation clinic discharge. If it should be the aim of the SMWC in the future to predict actual return to work rates better, there would be a need for a standardized checklist based on an evidence-based prediction model. Most German physicians conducting SMWCs in clinical routine at least partly agree with this statement and would be willing to use it if it does not take more than 10 minutes to apply. Future studies should develop and validate such a checklist. Once implemented in the German rehabilitation setting, it could improve the consistency and quality of assessments, potentially leading to better accuracy of the reality of return to work of patients with CRC after rehabilitation discharge.

## Data Availability

The data will not be published to an open access platform. After completion of the study, interested scientists can request data use and receive pseudonymized data upon approval of this application by the principal investigator. Please contact Dr. Ben Schoettker.

## Acknowledgements

We gratefully acknowledge the collaboration and commitment of all staff in the rehabilitation clinics involved in the MIRANDA study. We thank the documentarians at the German Cancer Research Centers’ Division of Clinical Epidemiology and Ageing Research for their work in the planning of data collection and all documentation procedures. We also thank all participants of the physician survey for their expert opinion given without any financial incentives.

## Contributions

Conceptualization, B.S.; methodology, T.V. and B.S.; validation, B.S. and T.V.; formal analysis, T.V. and B.S.; investigation, R.C., H.F., T.T., and M.S.; data curation, B.S.; writing—original draft preparation, T.V. and B.S.; writing—review and editing, T.V., B.S., R.C., H.F., T.T., M.S., and H.B.; visualization, T.V., B.S., and R.C.; supervision, B.S. and R.C.; project administration, M.S.; funding acquisition, H.B. and B.S. All authors have read and agreed to the published version of the manuscript.

## Ethics declarations

The study was conducted in accordance with the Declaration of Helsinki and in accordance with all applicable legal and regulatory requirements in Germany. The study was approved by the responsible Ethical Committee of the Faculty of Medicine Heidelberg (ethical approval code S-905/2019, date of approval: January 27 2020).

